# Quantitative Flow Ratio to Predict Non-Target-Vessel Events Prior to Planned Staged PCI in ACS Patients

**DOI:** 10.1101/2023.07.24.23292979

**Authors:** Sarah Bär, Raminta Kavaliauskaite, Tatsuhiko Otsuka, Yasushi Ueki, Jonas Häner, Jonas Lanz, Monika Fürholz, Fabien Praz, Lukas Hunziker, George CM Siontis, Thomas Pilgrim, Stefan Stortecky, Sylvain Losdat, Stephan Windecker, Lorenz Räber

## Abstract

**Background:** The optimal timepoint of staged percutaneous coronary intervention (sPCI) among patients with acute coronary syndrome (ACS) and multivessel disease (MVD) remains a matter of debate. Quantitative Flow Ratio (QFR) is a novel non-invasive method to assess the hemodynamic significance of coronary stenoses. We aimed to investigate whether QFR could optimize the timing of sPCI of non-target-vessels (non-TV) among ACS patients with MVD.

**Methods:** For this cohort study, ACS patients discharged from Bern University Hospital, Switzerland, were eligible if non-TV sPCI was scheduled within 6 months after index PCI. The primary endpoint was non-TV myocardial infarction (MI) and urgent unplanned non-TV PCI before planned sPCI. The association between lowest QFR per patient measured in non-TV (from index angiogram) and the primary endpoint was assessed using a multivariable adjusted Cox proportional hazards regression with QFR included as linear and penalized spline (non-linear) terms.

**Results:** QFR was measured in 1093/1432 ACS patients scheduled to undergo non-TV sPCI. Median time to sPCI was 28 days. The primary endpoint occurred in 5% of the patients. In multivariable analysis (1018 patients), there was no independent association between non-TV QFR and the primary endpoint (HR 0.87, 95% CI 0.69-1.05 [per 0.1 increase], p=0.125; non-linear p=0.648).

**Conclusions:** In ACS patients scheduled to undergo sPCI at a median of 4 weeks after index PCI, QFR did not emerge as independent predictor of non-TV events prior to planned sPCI. Thus, this study does not provide conceptual evidence, that QFR is helpful to optimize the timing of sPCI.

**Clinical Trial Registration:** ClinicalTrials.gov:NCT02241291

**Clinical Perspective:** **What is new?**

- This was the first study to investigate the association between non-target-vessel (non-TV) Quantitative Flow Ratio (QFR) and non-TV events occurring prior to planned staged percutaneous coronary intervention (PCI) among acute coronary syndrome (ACS) patients with multivessel disease, to derive first conceptual knowledge, whether QFR could be helpful to optimize the timing of staged PCI.
- Among 1093 ACS patients and 1262 non-TV scheduled to undergo out-of-hospital staged PCI within a median of 28 days from index PCI, QFR did not emerge as an independent predictor of non-TV events occurring prior to planned staged PCI.

**What are the clinical implications?**

- Among ACS patients in whom, according to the operator’s judgment, it is feasible to perform out-of-hospital staged PCI within a median of 1 month from index PCI, this study does not provide conceptual evidence, that QFR could be helpful to optimize the timing of staged PCI (i.e. to schedule staged PCI earlier in case of lower QFR).

**Grapical Abstract:** 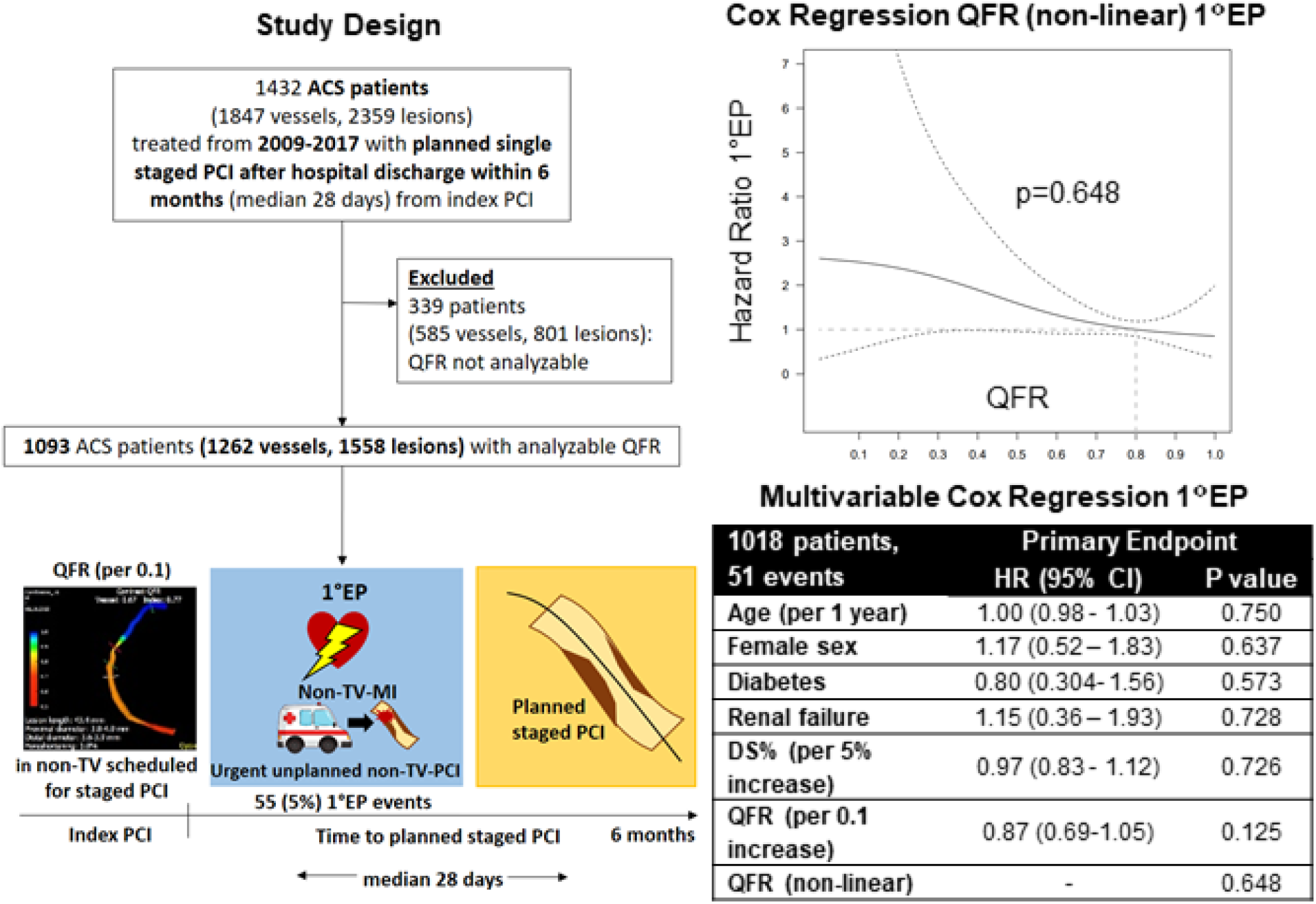

Study design (left) and primary endpoint results (right). For non-linear QFR, hazard ratios were calculated using the reference hazard corresponding to QFR=0.80 (grey dashed line) from a Cox proportional hazards model with penalized splines. ACS = acute coronary syndrome, CI = confidence interval, DS% = diameter stenosis, HR = hazard ratio, non-TV-MI = non-target-vessel myocardial infarction, non-TV-PCI = non-target-vessel percutaneous coronary intervention, PCI = percutaneous coronary intervention, QFR = Quantitative Flow Ratio, 1°EP = primary endpoint.

## INTRODUCTION

Multivessel disease (MVD) among patients with acute coronary syndromes (ACS) is present in up to 50% of patients with ST-segment elevation myocardial infarction (STEMI)^1^ and 40-80% with non-ST-elevation acute coronary syndrome (NSTE-ACS)^2^ and is associated with impaired prognosis.^3^ Complete revascularization results in improved clinical outcomes compared to culprit-lesion only revascularization in STEMI and NSTE-ACS.^4, 5^ Accordingly, complete revascularization obtained a Class IIa (level of evidence A^1^/C^6^) recommendation in current European Society of Cardiology (ESC) guidelines.^1, 6^ However, the optimal timepoint of non-target-vessel (non-TV) staged PCI remains a matter of ongoing debate.^1, 6, 7^ Coronary hemodynamics as assessed by Fractional Flow Reserve (FFR) of medically-treated non-culprit lesions have been reported to be inversely related to an increased risk of subsequent events among patients presenting with ACS.^8, 9^ But, it remains unknown, whether coronary hemodynamics are useful to determine the optimal timepoint of staged PCI in patients with ACS and MVD.

Fractional Flow Reserve (FFR) represents the current gold standard for the hemodynamic assessment of coronary lesions.^10^ Notwithstanding, FFR is infrequently used in patients with ACS owing to cost considerations, the invasive nature of the investigation, the need for vasodilator administration and the additional time required to complete the study.^11^ Quantitative Flow Ratio (QFR) is a novel, non-invasive, hyperemia-free method to calculate FFR derived from biplane coronary angiography using 3D-Quantitative Coronary Analysis (3D-QCA) and TIMI (Thrombolysis in Myocardial Infarction) frame counting.^12^ Among patients with chronic coronary syndrome (CCS), a QFR-based revascularization strategy using 0.80 as cut-off for ischemia (and treatment), has shown to improve 1-year clinical outcomes as compared to an angiography-guided approach.^13^ In ACS, good correlation with FFR^14^, predictive ability for clinical events^14, 15^, as well as good agreement between acute QFR compared to staged QFR have been reported.^14, 16, 17^

In order to assess whether QFR is able to determine the optimal timepoint of staged PCI in patients with ACS and MVD, we investigated the association between QFR of non-target vessels (non-TV) planned for staged PCI and non-TV events prior to planned staged PCI within the large, prospective Cardiobase Bern PCI registry.

## METHODS

### Patient Population

The Cardiobase Bern PCI registry (NCT02241291) is a prospective, single-center observational, registry of all consecutive patients undergoing PCI at Bern University Hospital, Switzerland established in 2009. There are no exclusion criteria other than inability or unwillingness to provide written informed consent. Baseline clinical, procedural and clinical outcomes are assessed at hospital discharge and 1 year after PCI by an independent clinical events committee. The registry complies with the Declaration of Helsinki and is approved by the institutional ethics committee.

Specific clinical in-and exclusion criteria for this investigation have been reported previously.^18^ In brief, all ACS patients included into the Cardiobase Bern PCI registry scheduled to undergo single staged PCI within 6 months from index PCI between 2009 and 2017 were eligible for this analysis. Patients with cardiogenic shock, in-hospital staged PCI, multiple staged PCIs, staged cardiac surgery, or missing information on staged PCI were excluded.^18^

### QFR and 3D-Quantitative Coronaray Angiography Analysis

QFR was assessed using the index procedure angiogram in the non-TV planned for staged PCI by experienced and certified analysts blinded for patient outcomes at the Corelab of Bern University Hospital, Switzerland using a dedicated software (QAngio XA 3D version 1.2, Medis Medical Imaging Systems, Leiden, the Netherlands). QFR specific exclusion criteria were absence of 2 projections with angle ≥25° apart, lack of isocenter calibration, substantial vessel overlap or vessel foreshortening, severe tortuosity, poor contrast, ostial left main or right coronary artery (RCA) stenosis, slow flow, tachycardia >100/min, and arrhythmias (i.e. atrial fibrillation, atrial flutter, idioventicular rhythm, (non-sustained) ventricular tachycardia). Contrast QFR using frame-counting^19^ was measured from the ostium to a distal landmark at a minimum of 1.5 mm distal vessel reference diameter, as reported previously.^15, 16^ The conventional QFR cut-off of ≤0.80 was used to detect significant ischemia.^12^ Lesion complexity was assessed according to the American College of Cardiology/American Heart Association (ACC/AHA) criteria.^20^ SB and RK had full access to all data and take responsibility for its integrity and the data analysis.

### Treatment

PCI was performed according to the recommendations and guidelines^1, 2, 6, 21^ valid at the time of presentation. Briefly, unfractionated heparin (initial bolus of 70-100 I.U. per kg body weight) was administrated during the procedure. Dual antiplatelet therapy (DAPT) consisting of acetylsalicylic acid and a potent P2Y12 inhibitor was initiated before or immediately after the index procedure. The recommended DAPT duration was usually 12 months from index treatment, but modified among patients taking oral anticoagulants or at high bleeding risk. Drug-eluting stents were routinely used. Staged procedures were usually performed between 3-8 weeks following index PCI according to institutional practice, but the exact timing was left to the operators’ discretion.

### Patient Follow-up

Patients were systematically and prospectively followed throughout 1 year to assess clinical outcomes and status of medical treatment. A health questionnaire was sent to all living patients with questions on re-hospitalization and adverse events, followed by telephone contact in case of missing response. General practitioners, referring cardiologists, and patients were contacted as necessary for additional information. For patients who underwent treatment for adverse events at other medical institutions, external medical records, discharge letters, and coronary angiography documentation were systematically collected and reviewed.

### Primary Analysis and Endpoint Definition

The primary analysis was an independent predictor analysis of the association between lowest QFR per patient (per 0.1 increase) and the composite of non-target vessel myocardial infarction (non-TV-MI) and urgent unplanned non-target-vessel PCI (non-TV-PCI), occurring before the planned staged PCI. MI was defined according to a modified historical definition.^22^ Non-target-vessel MI (non-TV-MI) was defined as MI attributed to non-culprit-vessels at baseline. Urgent unplanned non-TV-PCI was defined as urgent PCI in non-target-vessels performed earlier than planned due to ≥1 of the following: 1) recurrent MI^22^, 2) unstable angina^6^, 3) worsening congestive heart failure, 4) cardiogenic shock, or 5) symptomatic arrhythmia refractory to medication. This event had to be clearly distinguishable from the staged PCI procedure scheduled at index presentation.^18^ Clinical events were adjudicated by a clinical event committee consisting of two cardiologists (and a third one in case of disagreement) with use of original source documents.

Based on previous evidence on inverse, non-linear relationship between FFR and non-TV events plateauing at FFR 0.60^9^, we included a non-linear term for QFR. Other covariates were added based on clinical reasoning and consisted of age (per 1 year increase), female sex, renal failure (i.e. glomerular filtration rate (GFR) <60 ml/min), diabetes mellitus, and 3D-Quantitative Coronary Angiography (3D-QCA) diameter stenosis (DS%) (per 5% increase). In addition, we planned 2 sensitivity analyses:1) patient-level, using the same model as described, but with DS% replaced by ACC/AHA lesion complexity, and 2): vessel-level, using the following QFR and angiographic/3D-QCA characteristics: lowest QFR per patient (per 0.1 increase), 3D-QCA DS% (per 5% increase), minimum lumen diameter (MLD) (per 1mm increase), residual QFR (per 0.1 increase) (i.e. the residual QFR after virtual PCI predicted by an inherent algorithm in the QAngio XA 1.2 software), and ACC/AHA lesion complexity.

### Statistical Analysis

Continuous variables are expressed as mean ± standard deviation (SD) and categorical variables are expressed as counts with percentages. For the primary endpoint, we fitted univariable and multivariable Cox proportional hazards regressions including the following variables: lowest QFR per patient (per 0.1 increase) as linear and non-linear using penalized splines, age (per 1 year increase), female sex, renal failure, diabetes mellitus, and DS% (per 5% increase). As a sensitivity analysis for the primary endpoint we fitted the same models except that DS% was replaced by ACC/AHA lesion complexity. As a sensitivity analysis for the primary endpoint we also ran analyses at vessel-level. We used mixed-effects Cox proportional hazards models including patient identity as random factor to correct for multiple vessels per patient. Owing to the model’s higher complexity, we did not include a non-linear term for QFR. The variables in the model were lowest QFR per patient (per 0.1 increase), DS% (per 5% increase), MLD (per 1mm increase), residual QFR (per 0.1 increase), and ACC/AHA lesion complexity. For all multivariable models, we checked for the presence of multicollinearity by calculating the variance inflation factors (VIF) of all independent variables and confirmed that all VIFs were below 2.

Patients were censored at the time of the primary endpoint event, or at the time of the planned staged PCI, whichever occurred first. For vessel-level analysis, the culprit-vessel of a non-TV-MI was attributed a non-TV-MI event and an urgent unplanned non-TV PCI. If other vessels were treated during this same procedure, these were not adjudicated to have an event, since this treatment is likely to have been driven by logistical reasons, i.e. if a patient presents for another urgent invasive procedure, all remaining vessels are usually treated, even though they may not be responsible for the acute presentation. For urgent unplanned PCI, if no clear culprit-vessel could be identified from source data, all vessels treated during this procedure were adjudicated as urgent unplanned PCI event. Significance tests were two-tailed with a significance level set to 0.05. All analyses were performed using Stata 16 and R version 4.2.0.

## RESULTS

### Patient Population

Between January 2009 and December 2017, 8657 ACS patients (STEMI and NSTE-ACS) were consecutively enrolled into the Cardiobase Bern PCI Registry. Staged PCI was scheduled for 1764 patients of whom 1432 patients (1702 vessels, 2197 lesions) fulfilled the clinical eligibility criteria^18^ and were evaluated for QFR measurements. A total of 1262 vessels with 1558 lesions from 1093 patients were analyzable by QFR. Most frequent exclusion criteria were absence of 2 appropriate projections, missing angiographic data, or missing isocenter calibration (**Figure 1**). Baseline clinical characteristics and medical treatment at hospital discharge are summarized in **Table 1**. There was no significant differences in clinical characteristics between patients with QFR analysis available (N=1093) and those fulfilling the clinical eligibility criteria (N=1432)^18^ (**Table S1**). Mean patient age was 65 years, 88% were male, 17% had diabetes mellitus, 56% of patients presented with STEMI and 44% with NSTE-ACS. Median duration to planned staged PCI was 28 days (interquartile range (IQR) 28-42 days). Procedural characteristics of planned staged PCI and urgent unplanned non-TV-PCI are provided in **Table S2**.

**Figure 1.**
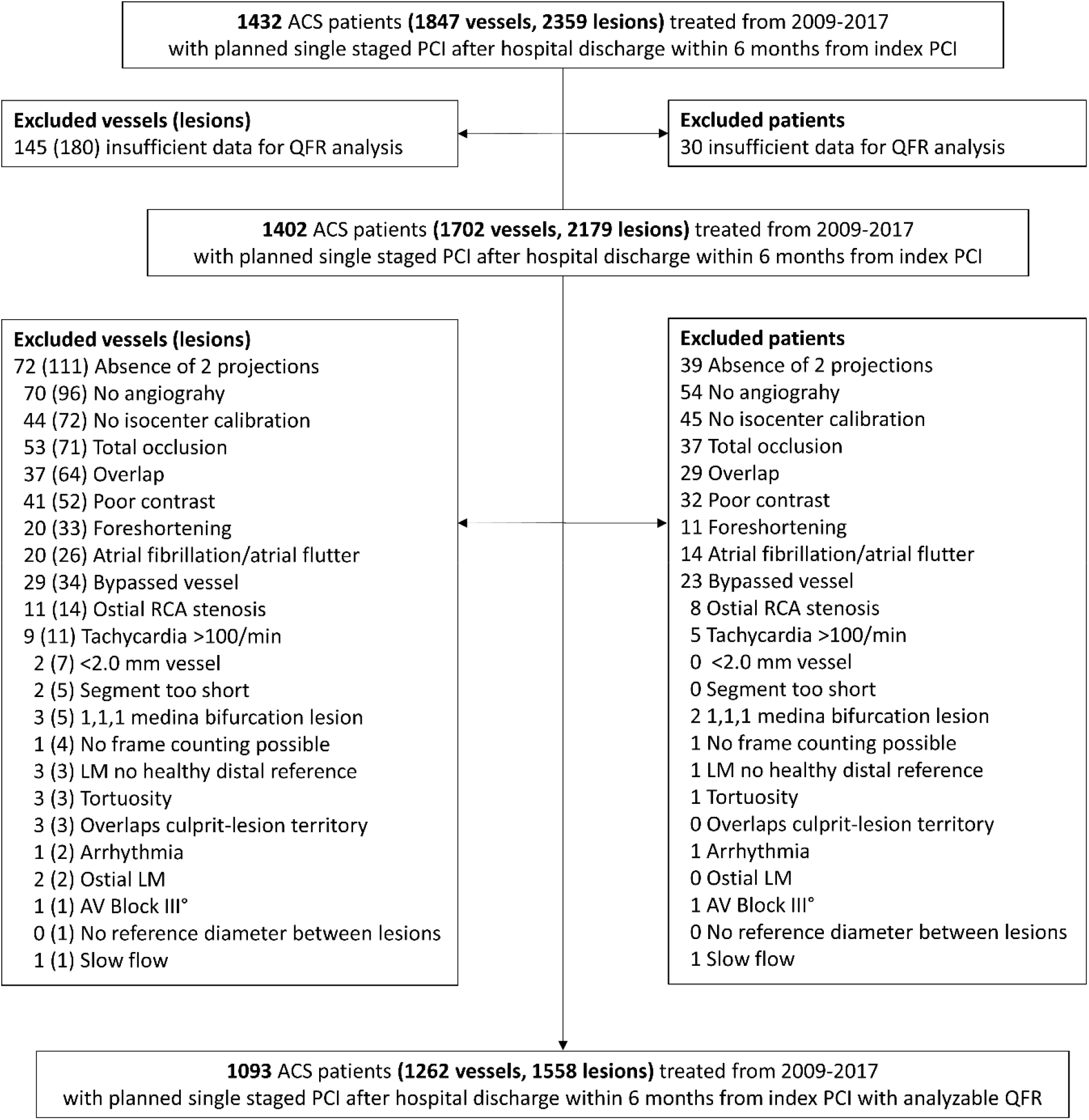
Flowchart. ACS = acute coronary syndrome, LM = left main, PCI = percutaneous coronary intervention, QFR = Quantitative Flow Ratio, RCA = right coronary artery.

**Table 1.** Patient Characteristics.

|  | **Patients**<br>**N=1093** |
| --- | --- |
| Age, years | 65 ±11 |
| Female, n (%) | 238 (22%) |
| BMI, kg/m<sup>2</sup> | 27 ±4.2 |
| Smoker, n (%) | 403 (37%) |
| Hypercholesterolemia, n (%) | 564 (52%) |
| Hypertension, n (%) | 633 (58%) |
| Diabetes mellitus, n (%) | 188 (17%) |
| Family history of CAD, n (%) | 267 (24%) |
| Previous MI, n (%) | 62 (5.7%) |
| Previous PCI, n (%) | 92 (8.4%) |
| Previous CABG, n (%) | 12 (1.1%) |
| Left ventricular function, % | 51 ±11 |
| Indication, n (%) |  |
| Unstable Angina | 45 (4.1%) |
| NSTEMI | 440 (40%) |
| STEMI | 608 (56%) |
| Congestive heart failure, n (%) |  |
| Killip I | 948 (87%) |
| Killip II | 113 (10%) |
| Killip III | 32 (2.9%) |
| Renal failure (GFR <60 ml/min), n (%) | 172 (16%) |
| Renal failure requiring dialysis, n (%) | 15 (1.4%) |
| Peripheral arterial disease, n (%) | 42 (3.8%) |
| History of stroke or TIA, n (%) | 46 (4.2%) |
| History of GI bleeding, n (%) | 13 (1.2%) |
| History of malignancy, n (%) | 93 (8.5%) |
| COPD, n% | 57 (5.2%) |
| Anemia<sup>*</sup>, n (%) | 155 (14%) |
| Days from index to planned staged PCI | 28 [28, 42] |
| **Medication at hospital discharge** |  |
| Aspirin, n (%) | 1082 (99%) |
| Potent P2Y12 inhibitor (Prasugrel or Ticagrelor), n (%) | 851 (78%) |
| Clopidogrel, n (%) | 240 (22%) |
| Any DAPT, n (%) | 1081 (99%) |
| Oral anticoagulation (vitamin K antagonists or NOAC), n (%) | 60 (5.5%) |
| Statin, n (%) | 1043 (95%) |

Values are n (%), mean ± standard deviations (SD), or median [interquartile range (IQR)]. <sup>\*</sup> Anemia was defined as hemoglobin <130 g/l in men and <120 g/l in women. ACS = acute coronary syndrome, BMI = body mass index, CABG = coronary artery bypass graft, CAD = coronary artery disease, COPD = chronic obstructive pulmonary disease (COPD), DAPT = dual antiplatelet therapy, GFR = glomerular filtration rate, GI = gastrointestinal, NOAC = novel oral anticoagulant, NSTEMI = non-ST-segment-elevation myocardial infarction, PCI = percutaneous coronary intervention, STEMI = ST-elevation-segment myocardial infarction, TIA = transitory ischemic attack.

### QFR and 3D-QCA Characteristics

Of 1262 vessels analyzed by QFR, 41.1% were left anterior descending (LAD) (n=519), 30.6% left circumflex (LCX) (n=386), 27.1% RCA (n=342), and 1.2% (n=15) left main (LM) vessels. Mean QFR per patient was QFR 0.73±0.17, mean DS% 54.8 ±11.2%, and ACC/AHA angiographic lesion complexity (lesion-level) was most frequently C, followed by B1 (**Table 2**, **Figure 2**).

**Figure 2.**
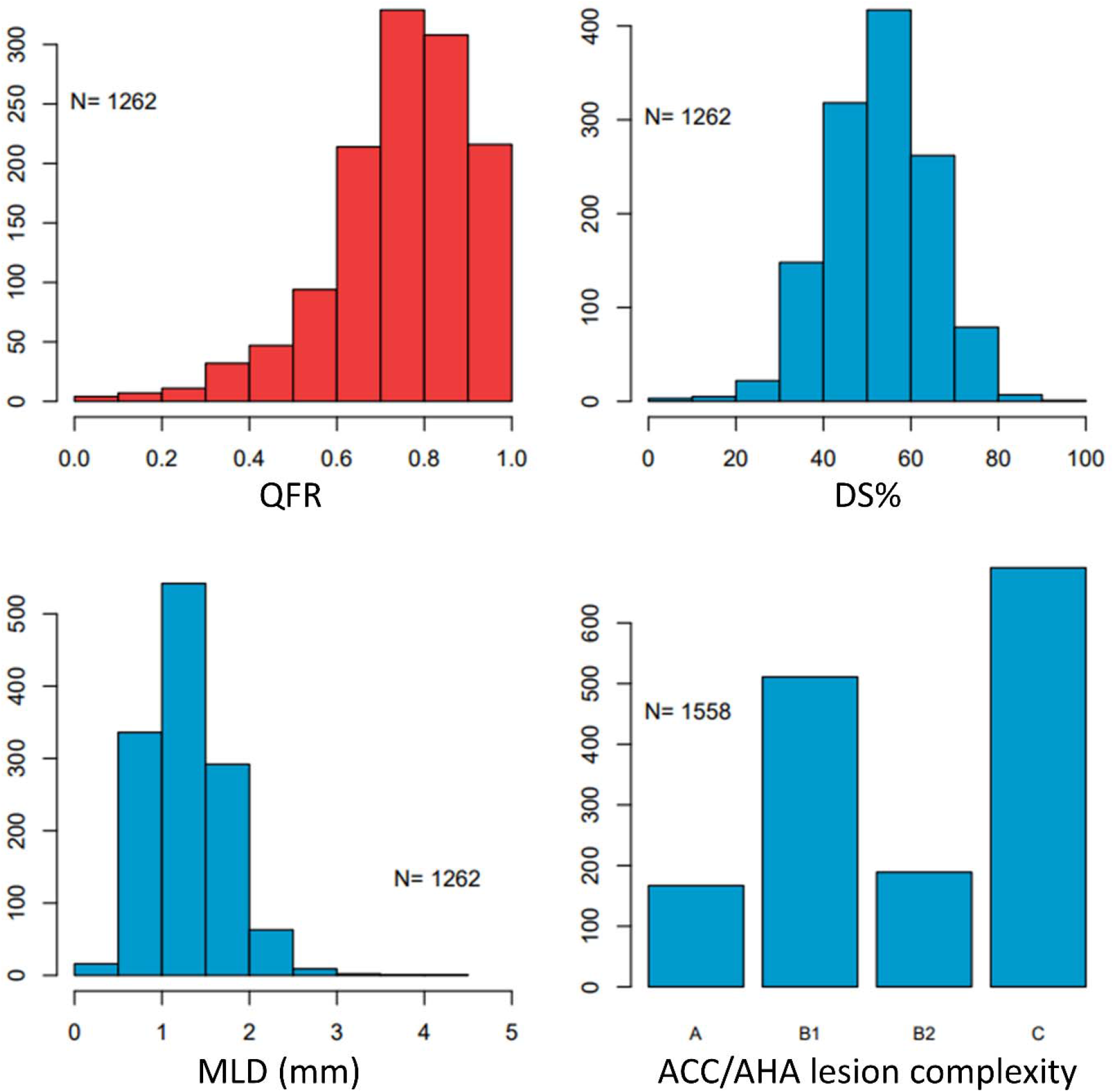
QFR and 3D-QCA Characteristics. ACC/AHA = American College of Cardiology/American Heart Association, DS% = diameter stenosis, MLD = minimum lumen diameter, QFR = Quantitative Flow Ratio, 3D-QCA =3D-Quantitative Coronary Angiography.

**Table 2.** QFR and 3D-QCA.

|  | **Patients (N=1093)** | **Vessels (N=1262)** |
| --- | --- | --- |
| Diameter stenosis, % | 54.8 ±11.2 | 53.6 ±11.5 |
| Area stenosis, % | 70.4 ±12.6 | 69.0 ±13.4 |
| Lesion length, mm | 26.1 ±12.1 | 25.0 ±12.0 |
| Proximal diameter, mm | 2.86 ±0.59 | 2.82 ±0.59 |
| Distal diameter, mm | 2.45 ±0.57 | 2.47 ±0.57 |
| Minimum lumen diameter, mm | 1.36 ±0.57 | 1.34 ±0.56 |
| QFR | 0.73 ±0.17 | 0.75 ±0.17 |

Values are mean ±standard deviation (SD). QFR = Quantitative Flow Ratio.
3D-QCA = 3D-Quantitative Coronary Angiography.

### Primary and Secondary Analyses

A total 55 (5.0%) of primary endpoint events had occurred within median 11 days (interquartile range (IQR) 5-16 days) prior to planned staged PCI. In multivariable analysis (1018 patients 51 events), there was no independent association between QFR and the primary endpoint (HR 0.87, 95% confidence interval (CI) 0.69-1.05, p=0.125, QFR non-linear p=0.648) (**Graphical Abstract, Table 3**). Overall, none of the variables in the model showed a significant association with the clinical outcomes (**Table 3**). The sensitivity analysis on patient-level using ACC/AHA lesion complexity instead of DS% showed consistent results (**Table 3**). Also, in the sensitivity analysis on vessel-level, there was no independent association between QFR and the primary endpoint (multivariable HR 0.84, 95% CI 0.65-1.04), p=0.083) (**Table 3**). Cumulative event curves of the primary endpoint components and planned staged PCI are shown in **Figure 3**.

**Figure 3.**
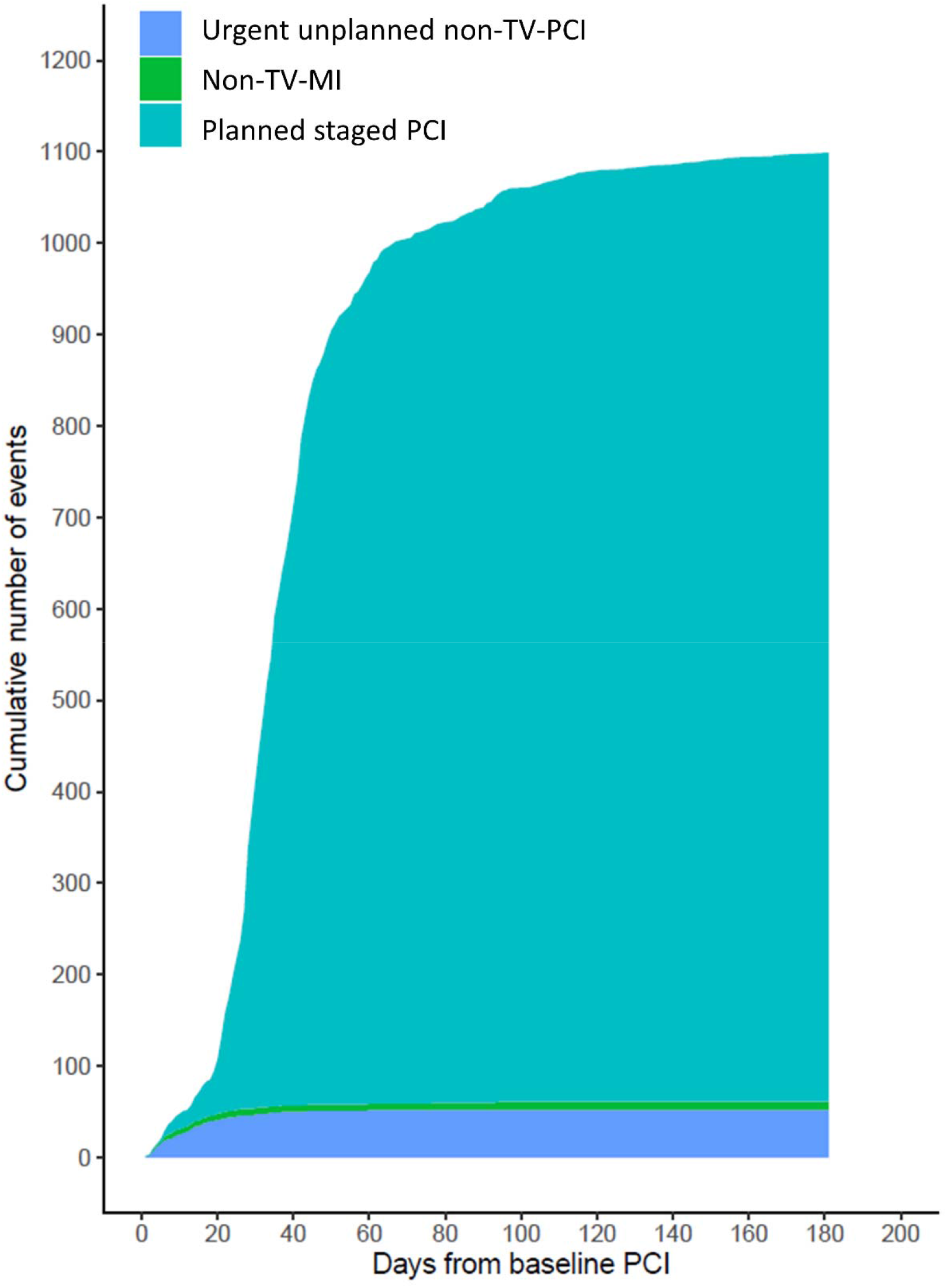
Cumulative Event Curves. Cumulative event curves of urgent unplanned non-target-vessel (non-TV) staged percutaneous coronary intervention (PCI), non-TV myocardial infarction (MI), and planned staged PCI.

**Table 3.**
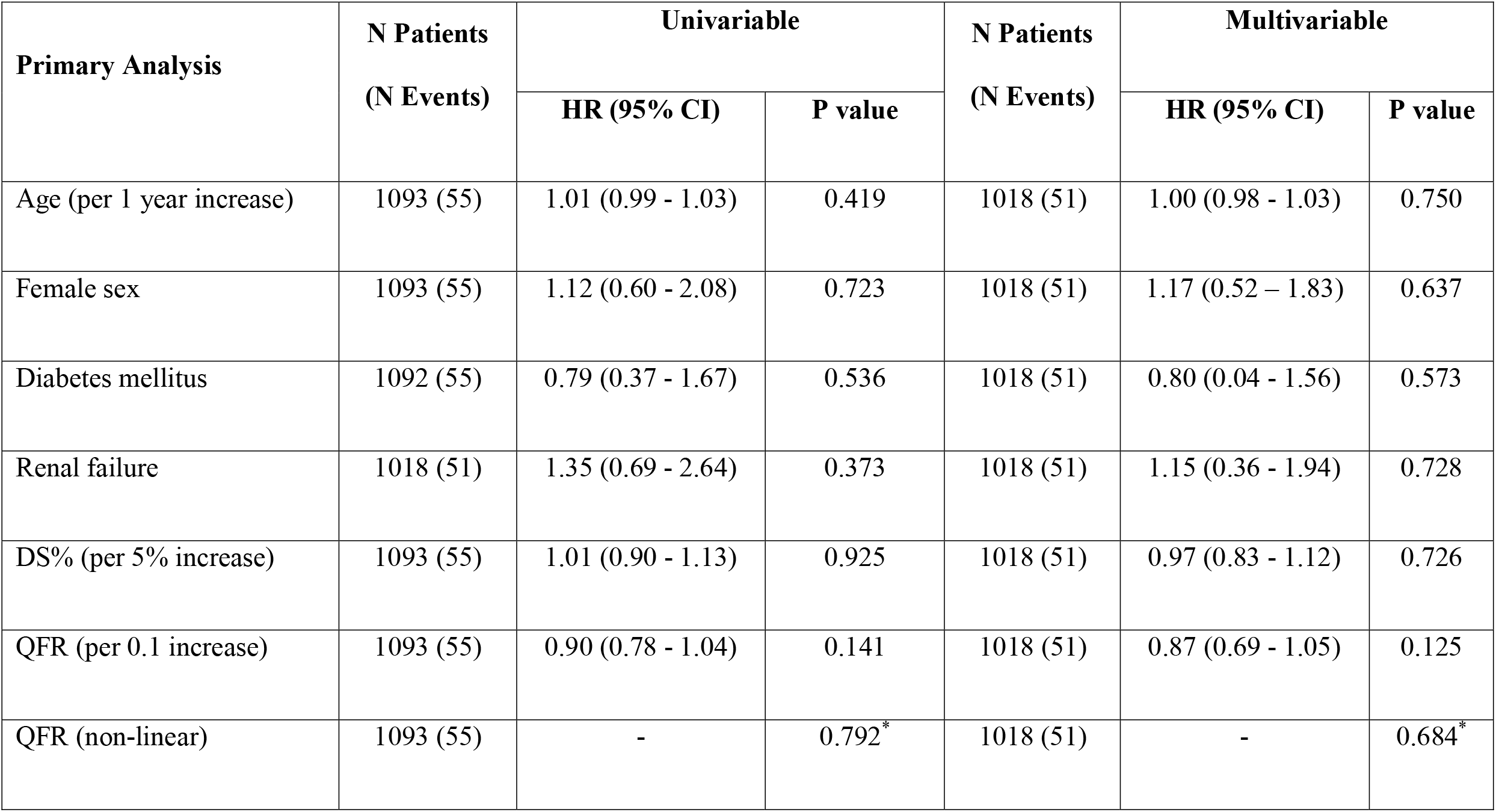
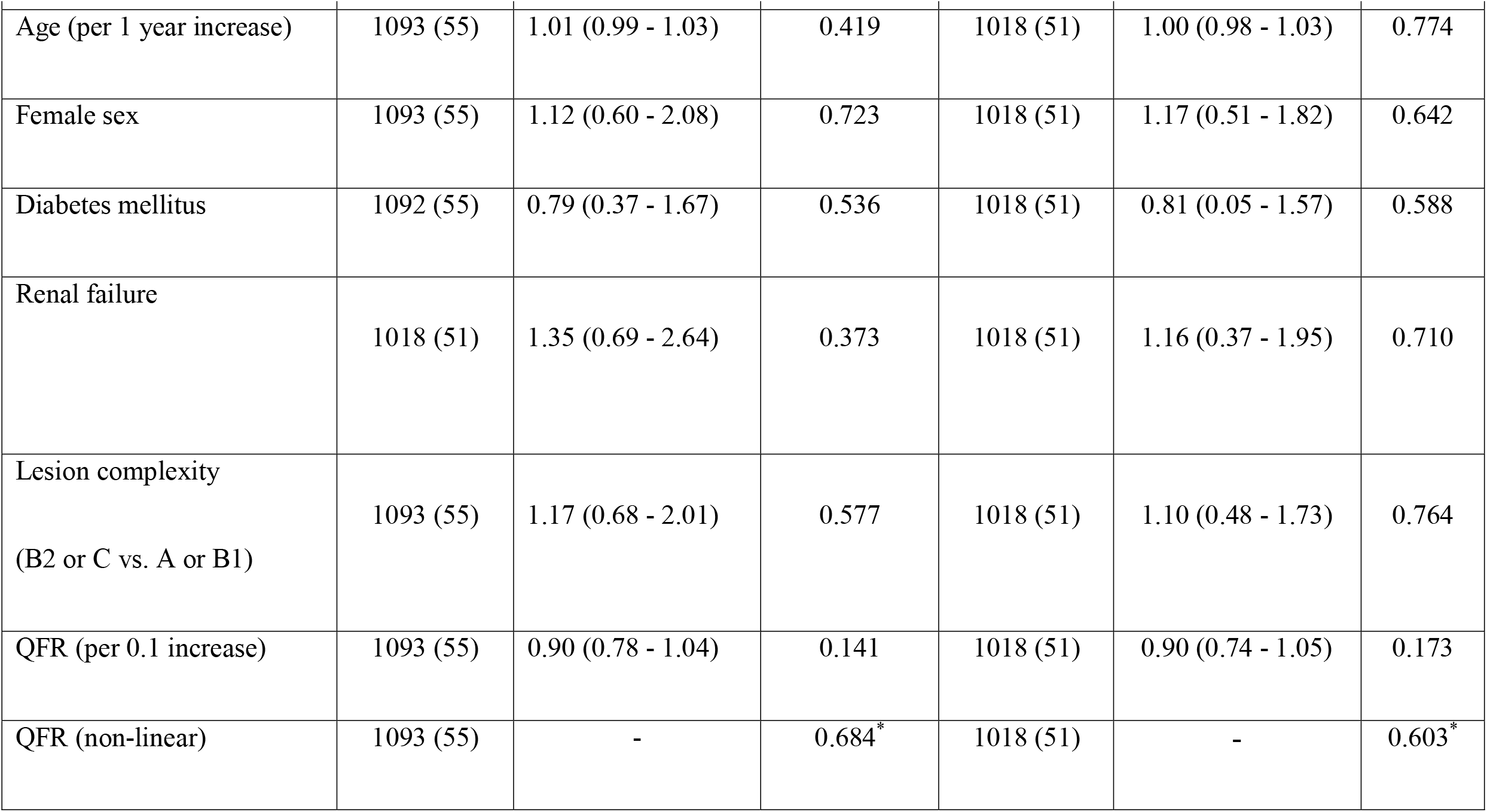
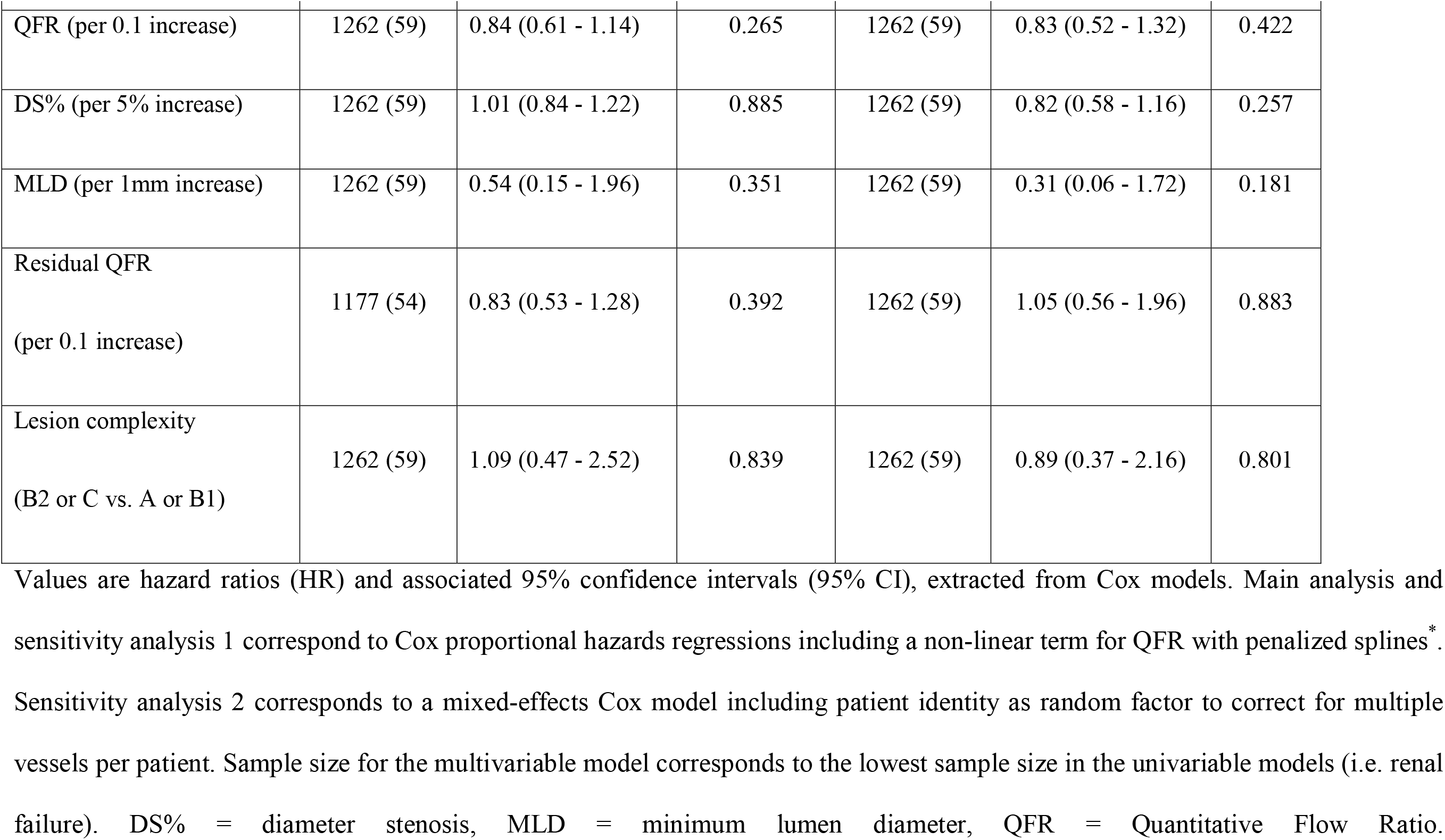
Cox Regressions Primary Endpoint.

| Primary Analysis | N Patients<br>(N Events) | Univariable |  | N Patients<br>(N Events) | Multivariable |  |
| --- | --- | --- | --- | --- | --- | --- |
|  |  | HR (95% CI) | P value |  | HR (95% CI) | P value |
| Age (per 1 year increase) | 1093 (55) | 1.01 (0.99 - 1.03) | 0.419 | 1018 (51) | 1.00 (0.98 - 1.03) | 0.750 |
| Female sex | 1093 (55) | 1.12 (0.60 - 2.08) | 0.723 | 1018 (51) | 1.17 (0.52 – 1.83) | 0.637 |
| Diabetes mellitus | 1092 (55) | 0.79 (0.37 - 1.67) | 0.536 | 1018 (51) | 0.80 (0.04 - 1.56) | 0.573 |
| Renal failure | 1018 (51) | 1.35 (0.69 - 2.64) | 0.373 | 1018 (51) | 1.15 (0.36 - 1.94) | 0.728 |
| DS% (per 5% increase) | 1093 (55) | 1.01 (0.90 - 1.13) | 0.925 | 1018 (51) | 0.97 (0.83 - 1.12) | 0.726 |
| QFR (per 0.1 increase) | 1093 (55) | 0.90 (0.78 - 1.04) | 0.141 | 1018 (51) | 0.87 (0.69 - 1.05) | 0.125 |
| QFR (non-linear) | 1093 (55) | - | 0.792<sup>*</sup> | 1018 (51) | - | 0.684<sup>*</sup> |

| **Sensitivity Analysis 1** | **N Patients**<br>**(N Events)** | **Univariable** |  | **N Patients**<br>**(N Events)** | **Multivariable** |  |
| --- | --- | --- | --- | --- | --- | --- |
|  |  | **HR (95% CI)** | **P value** |  | **HR (95% CI)** | **P value** |
| Age (per 1 year increase) | 1093 (55) | 1.01 (0.99 - 1.03) | 0.419 | 1018 (51) | 1.00 (0.98 - 1.03) | 0.774 |
| Female sex | 1093 (55) | 1.12 (0.60 - 2.08) | 0.723 | 1018 (51) | 1.17 (0.51 - 1.82) | 0.642 |
| Diabetes mellitus | 1092 (55) | 0.79 (0.37 - 1.67) | 0.536 | 1018 (51) | 0.81 (0.05 - 1.57) | 0.588 |
| Renal failure | 1018 (51) | 1.35 (0.69 - 2.64) | 0.373 | 1018 (51) | 1.16 (0.37 - 1.95) | 0.710 |
| Lesion complexity<br>(B2 or C vs. A or B1) | 1093 (55) | 1.17 (0.68 - 2.01) | 0.577 | 1018 (51) | 1.10 (0.48 - 1.73) | 0.764 |
| QFR (per 0.1 increase) | 1093 (55) | 0.90 (0.78 - 1.04) | 0.141 | 1018 (51) | 0.90 (0.74 - 1.05) | 0.173 |
| QFR (non-linear) | 1093 (55) | - | 0.684<sup>*</sup> | 1018 (51) | - | 0.603<sup>*</sup> |

| **Sensitivity Analysis 2** | **N Vessels (N**<br>**Events)** | **Univariable** |  | **N Vessels (N**<br>**Events)** | **Multivariable** |  |
| --- | --- | --- | --- | --- | --- | --- |
|  |  | **HR (95% CI)** | **P value** |  | **HR (95% CI)** | **P value** |
| QFR (per 0.1 increase) | 1262 (59) | 0.84 (0.61 - 1.14) | 0.265 | 1262 (59) | 0.83 (0.52 - 1.32) | 0.422 |
| DS% (per 5% increase) | 1262 (59) | 1.01 (0.84 - 1.22) | 0.885 | 1262 (59) | 0.82 (0.58 - 1.16) | 0.257 |
| MLD (per 1mm increase) | 1262 (59) | 0.54 (0.15 - 1.96) | 0.351 | 1262 (59) | 0.31 (0.06 - 1.72) | 0.181 |
| Residual QFR<br>(per 0.1 increase) | 1177 (54) | 0.83 (0.53 - 1.28) | 0.392 | 1262 (59) | 1.05 (0.56 - 1.96) | 0.883 |
| Lesion complexity<br>(B2 or C vs. A or B1) | 1262 (59) | 1.09 (0.47 - 2.52) | 0.839 | 1262 (59) | 0.89 (0.37 - 2.16) | 0.801 |

Values are hazard ratios (HR) and associated 95% confidence intervals (95% CI), extracted from Cox models. Main analysis and sensitivity analysis 1 correspond to Cox proportional hazards regressions including a non-linear term for QFR with penalized splines<sup>\*</sup>. Sensitivity analysis 2 corresponds to a mixed-effects Cox model including patient identity as random factor to correct for multiple vessels per patient. Sample size for the multivariable model corresponds to the lowest sample size in the univariable models (i.e. renal failure). DS% = diameter stenosis, MLD = minimum lumen diameter, QFR = Quantitative Flow Ratio.

## DISCUSSION

In this cohort study of patients with ACS and MVD scheduled to undergo out-of-hospital staged PCI within a median of 28 (IQR 28-42) days from index presentation, non-TV QFR of vessels scheduled for staged PCI using the index procedure angiogram did not show an independent association with non-TV events occurring prior to the planned staged PCI. Therefore, this study does not provide conceptual evidence for QFR being able to optimize the timing of staged PCI (i.e. to plan earlier in case of lower QFR). These results apply to patients scheduled on average 4 weeks after the index procedure and mean QFR value of 0.73 in the untreated non-culprit vessel.

### Current Recommendations on Non-Culprit-Lesion Revascularization in ACS

Current ESC guidelines on the management of STEMI^1^ provide a Class IIa (level of evidence A) recommendation for non-culprit-lesion revascularization during the index procedure or before hospital discharge, based on the treatment strategies in the RCTs which compared culprit-lesion only vs. complete revascularization until the release of the guidelines.^23–26^ In the COMPLETE (*Complete versus Culprit-Only Revascularization Strategies to Treat Multivessel Disease after Early PCI for STEMI*) trial^5^, a large-scale RCT comparing complete revascularization vs. culprit-lesion only PCI in STEMI patients, the benefit of complete revascularization over culprit-lesion only PCI was independent of whether staged PCI was performed during index hospitalization (median 1 day, IQR 1-3 days) or after hospital discharge (median 23 days, IQR 12.5-33.5 days).^7^ In line, with this investigation, we had observed similar outcomes with early (i.e. <4 weeks from index PCI) vs. late (≥4 weeks from index PCI) staged PCI^18^ in the same study population as reported here.

For NSTE-ACS, a Class IIb (level of evidence B) recommendation is given for immediate complete revascularization based on the RCT SMILE^27^ (*Impact of Different Treatment in Multivessel Non ST Elevation Myocardial Infarction Patients: One Stage Versus Multistaged Percutaneous Coronary Intervention*), where immediate complete revascularization was superior as compared to in-hospital staged PCI.^6^ However, more emphasis (Class I, level of evidence B) is given to individual lesion characteristics including the functional significance of non-culprit lesions.^6^

In the recently published RCT BIOVASC (*Percutaneous Complete Revascularization Strategies Using Sirolimus-Eluting Biodegradable Polymer-Coated Stents in Patients Presenting With Acute Coronary Syndrome and Multivessel Disease*)^28^ including 40% STEMI and 60% NSTE-ACS patients, immediate complete revascularization was non-inferior to staged complete revascularization in-hospital or within 6 weeks (median 15 days, IQR 4-28 days) from index presentation. Immediate complete revascularization was associated with a reduction in early recurrent myocardial infarction and unplanned ischemia-driven revascularization, and for 30-day outcomes, superiority for immediate complete revascularization was met for the primary endpoint (death, myocardial infarction, unplanned ischemia-driven revascularization, cerebrovascular events). While indicating, that immediate complete revascularization is safe (in hemodynamically stable patients) and potentially protective with respect to early events before planned staged PCI, no evidence on the optimal duration to staged PCI can be derived from this trial. Additional RCTs are currently investigating the issue (MULTISTARS AMI (NCT03135275), STAGED (NCT04918030), OPTION-NSTEMI (NCT04968808)).

### Link between Coronary Physiology and Non-TV-Events

In a subanalysis of the COMPARE-ACUTE (*Comparison Between FFR Guided Revascularization Versus Conventional Strategy in Acute STEMI Patients With MVD)* trial^9^, an inverse non-linear relationship between deferred lesions among STEMI patients investigated by FFR and non-TV events was observed which plateaued at FFR 0.60. Additional evidence exists from retrospective analyses from mixed populations including 29% ACS, where FFR was shown to be continuously and inversely related to ischemic event risk.^8^ These analyses support the concept that the functional significance of non-culprit lesions may represent an ischemic continuum with increasing inverse event risk, rather than a dichotomous state dividing at FFR 0.80. With respect to QFR, among patients with CCS, QFR-guided revascularization has been reported to improve 1-year outcomes as compared to angiography-guided revascularization^13^. Further, in STEMI patients, acute QFR shows even better agreement with 30-day FFR as acute FFR itself^29^, which in addition to its non-invasive and hyperemia-free nature, makes it an interesting diagnostic tool for the ACS population.

However, in our investigation, we did not observe any independent association between QFR and non-TV events occurring before staged PCI. These findings suggest that non-TV QFR is unable to refine the timing of staged PCI, among patients undergoing operators’ scheduled out-of-hospital staged PCI within a median of 28 days from index PCI. The overall event rate was 5%, the number of very low QFR values (i.e. <0.60) small, and the timeframe for the events to occur short with a median of 28 days. Therefore, we cannot definitely exclude a potential association between QFR and non-TV events prior to staged PCI in a larger patient population with more pronounced ischemia. Along these lines, we observed a trend towards higher clinical events with lower QFR, but this investigation is limited by low sample size (**Figure S1**). Interestingly, none of the other classical covariates in the prediction models showed an independent association with the primary endpoint, implying that other factors seem to drive this type of event.

### Plaque Morphology as a Potential Driver of Events

At variance to the clinical setting of CCS demonstrating improved short-and middle-term outcomes with physiology-guided compared to angiography-guided revascularization using FFR^30^ and QFR^13^, no superiority of FFR-guided vs. angiography-guided complete non-culprit lesion revascularization has been observed in the STEMI population.^31^ Vulnerable non-culprit plaque features such as high plaque burden, thin fibrous cap and low minimal lumen area are highly prevalent in ACS patients^32^ and have been shown to be associated with subsequent events.^32^ Therefore, and based on the findings of this study, it may be hypothesized, that plaque morphology may play a more important role as compared to physiology in driving early non-TV events occurring before planned staged PCI. This should be investigated in future intracoronary imaging studies.

### Limitations

The study results need to be considered in light of several limitations: 1) It is an observational, non-randomized, post-hoc, single-center study. 2) The timepoint of staged PCI (and thus time to event) was defined by operators’ judgement, however the aim of the study was to investigate potential add-on value of QFR for the timing of staged PCI on top of clinical judgement and not QFR alone. 3) The number of vessels with very low QFR was limited and we observed a trend towards a higher percentage of events with lower QFR, so that we cannot exclude a potential association between QFR and non-TV events occurring before staged PCI in larger patient populations with more pronounced ischemia. 4) Highest-risk patients, i.e. those undergoing in-hospital staged PCI (n=139) or those with cardiogenic shock at index presentation (n=70), were excluded from this study^18^, and thus, the results do not pertain to this higher risk population. 5) FFR and intracoronary imaging were used clinically upon the discretion of the operators and could not be collected systematically.

## Conclusions

In this cohort study of 1093 patients with ACS and MVD scheduled to undergo out-of-hospital staged PCI within 28 to 42 days from index presentation, non-TV QFR derived from vessels planned for staged PCI using the baseline angiogram was not associated with non-TV events prior to staged PCI. Therefore, this study does not provide conceptual evidence, that QFR is able to optimize the timing of staged PCI. The concept may warrant further investigation among larger populations with more pronounced ischemia and longer duration to staged PCI.

## Disclosures

Dr. Bär reports research grants to the institution from Medis Medical Imaging Systems, Abbott, Bangerter-Rhyner Stiftung, and a personal research grant from the Swiss National Science Foundation, outside the submitted work. Dr. Ueki reports personal fees from Infraredex, outside the submitted work. Dr. Losdat is employed by CTU Bern, University of Bern, which has a staff policy of not accepting honoraria or consultancy fees. However, CTU Bern is involved in design, conduct, or analysis of clinical studies funded by not-for-profit and for-profit organizations. In particular, pharmaceutical and medical device companies provide direct funding to some of these studies. For an up-to-date list of CTU Bern’s conflicts of interest: http://www.ctu.unibe.ch/research/declaration_of_interest/index_eng.html. Prof. Praz has received travel expenses from Abbott Vascular, Edwards Lifesciences, and Polares Medical. Dr. Häner has received a travel grant from Bayer. Prof. Stortecky reports research grants to the institution from Edwards Lifesciences, Medtronic, Abbott Vascular, and Boston Scientific; speaker fees from Boston Scientific; consulting fees from BTG and Teleflex, outside the submitted work. Prof. Pilgrim reports research grants to the institution from Boston Scientific, Biotronik, and Edwards Lifesciences; speaker fees from Boston Scientific and Biotronik; Consultancy for HighLife SAS; proctoring for Boston Scientific and Medtronic. Stephan Windecker reports research, travel or educational grants to the institution without personal remuneration from Abbott, Abiomed, Amgen, Astra Zeneca, Bayer, Braun, Biotronik, Boehringer Ingelheim, Boston Scientific, Bristol Myers Squibb, Cardinal Health, CardioValve, Cordis Medical, Corflow Therapeutics, CSL Behring, Daiichi Sankyo, Edwards Lifesciences, Farapulse Inc. Fumedica, Guerbet, Idorsia, Inari Medical, InfraRedx, Janssen-Cilag, Johnson & Johnson, Medalliance, Medicure, Medtronic, Merck Sharp & Dohm, Miracor Medical, Novartis, Novo Nordisk, Organon, OrPha Suisse, Pharming Tech. Pfizer, Polares, Regeneron, Sanofi-Aventis, Servier, Sinomed, Terumo, Vifor, V-Wave. Stephan Windecker served as advisory board member and/or member of the steering/executive group of trials funded by Abbott, Abiomed, Amgen, Astra Zeneca, Bayer, Boston Scientific, Biotronik, Bristol Myers Squibb, Edwards Lifesciences, MedAlliance, Medtronic, Novartis, Polares, Recardio, Sinomed, Terumo, and V-Wave with payments to the institution but no personal payments. He is also member of the steering/executive committee group of several investigator-initiated trials that receive funding by industry without impact on his personal remuneration. Prof. Räber reports research grants to the institution from Abbott Vascular, Biotronik, Boston Scientific, Heartflow, Sanofi, Regeneron, Medis Medical Imaging Systems, Bangerter-Rhyner Stiftung; speaker or consultation fees by Abbott Vascular, Amgen, AstraZeneca, Canon, Novo Nordisk, Medtronic, Occlutech, and Sanofi, outside the submitted work. All other authors report no conflicts of interest.

### Non-standard Abbreviations and Acronyms

MVD: multivessel disease
Non-TV: non-target vessel
QFR: Quantitative Flow Ratio
3D-QCA: 3D-Quantitative Coronary Angiography

## Data Availability

The data that support the findings of this study will be made available upon reasonable request from the corresponding author.

## Supplemental Material

Supplemental Tables S1-S2

Supplemental Figures S1-S2

