## Supplemental Material for "Quantitative Flow Ratio to Predict Non-Target-Vessel Events Prior to Planned Staged PCI in ACS Patients"

**Table S1. Patient Characteristics of QFR Cohort vs. Full Study Cohort[18]**

| | QFR Cohort<br>N=1093 | Full Cohort<br>N=1432 |
|---|---|---|
| Age, years | 65.2 ±11.4 | 65.5 ±11.4 |
| Female, n (%) | 238 (22%) | 300 (21%) |
| BMI, kg/m$^2$ | 27.3 ±4.2 | 27.3 ±4.3 |
| Smoker, n (%) | 403 (37%) | 524 (37%) |
| Hypercholesterolemia, n (%) | 564 (52%) | 754 (53%) |
| Hypertension, n (%) | 633 (58%) | 858 (60%) |
| Diabetes mellitus, n (%) | 188 (17%) | 274 (19%) |
| Family history of CAD, n (%) | 267 (24%) | 349 (24%) |
| Previous MI, n (%) | 62 (5.7%) | 106 (7.4%) |
| Previous PCI, n (%) | 92 (8.4%) | 142 (10%) |
| Previous CABG, n (%) | 12 (1.1%) | 40 (2.8%) |
| Left ventricular function, % | 50.7 ±11.1 | 49.9 ±11.5 |
| Indication, n (%) | | |
| Unstable Angina | 45 (4.1%) | 71 (5.0%) |
| NSTEMI | 440 (40%) | 558 (39%) |
| STEMI | 608 (56%) | 803 (56%) |
| Congestive heart failure, n (%) | | |
| Killip I | 948 (87%) | 1220 (85%) |
| Killip II | 113 (10%) | 158 (11%) |
| Killip III | 32 (2.9%) | 54 (3.8%) |

| | | |
|---|---|---|
| Renal failure (GFR <60 ml/min), n (%) | 172 (16%) | 224 (16%) |
| Renal failure requiring dialysis, n (%) | 15 (1.4%) | 19 (1.3%) |
| Peripheral arterial disease, n (%) | 42 (3.8%) | 58 (4.1%) |
| History of stroke or TIA, n (%) | 46 (4.2%) | 64 (4.5%) |
| History of GI bleeding, n (%) | 13 (1.2%) | 18 (1.3%) |
| History of malignancy, n (%) | 93 (8.5%) | 120 (8.4%) |
| COPD, n% | 57 (5.2%) | 75 (5.2%) |
| Anemia*, n (%) | 155 (14%) | 211 (15%) |
| Days from index to planned staged PCI | 28 [28, 42] | 28 [28, 42] |

Values are n (%), mean ± standard deviations (SD), or median [interquartile range (IQR)]. Full study cohort refers to reference 1. *Anemia was defined as hemoglobin <130 g/l in men and <120 g/l in women. ACS = acute coronary syndrome, BMI = body mass index, CABG = coronary artery bypass graft, CAD = coronary artery disease, COPD = chronic obstructive pulmonary disease (COPD), GFR = glomerular filtration rate, GI = gastrointestinal, NSTEMI = non-ST-segment-elevation myocardial infarction, PCI = percutaneous coronary intervention, STEMI = ST-elevation-segment myocardial infarction, TIA = transitory ischemic attack.

**Table S2. Procedural Characteristics of Staged PCI and Urgent Unplanned Non-TV-PCI**

| | Planned staged PCI | Urgent unplanned non-TV PCI |
|---|---|---|
| | **N=1041 Patients** | **N=52 Patients** |
| Number of vessels, n (%) | | |
| 1 | 879 (84%) | 42 (81%) |
| 2-3 | 162 (16%) | 10 (19%) |
| Number of lesions, n (%) | | |
| 1 | 589 (57%) | 32 (62%) |
| 2 | 313 (30%) | 13 (25%) |
| 3 | 112 (11%) | 5 (10%) |
| ≥4 | 27 (2.6%) | 2 (3.8%) |
| | **N=1476 Lesions** | **N=82 Lesions** |
| Target vessel, n (%) | | |
| Left main | 15 (1.0%) | 0 (0.0%) |
| Left anterior descending | 637 (43%) | 32 (39%) |
| Left circumflex | 420 (28%) | 31 (38%) |
| Right coronary artery | 404 (27%) | 19 (23%) |
| Type of intervention, n (%) | | |
| Implantation of stent(s) | 1422 (96%) | 78 (95%) |
| Balloon dilatation only | 53 (3.4%) | 4 (4.9%) |
| Escalation to CABG* | 5 (0.3%) | 0 (0.0%) |
| Restenotic lesion, n (%) | 11 (0.7%) | 0 (0.0%) |

| | | |
|---|---|---|
| Thrombus aspiration, n (%) | 2 (0.1%) | 0 (0.0%) |
| Total number of stents implanted, n | | |
| 1 | 1072 (73%) | 63 (77%) |
| 2 | 296 (20%) | 13 (16%) |
| ≥3 | 108 (7.3%) | 6 (7.3%) |
| Total stent length, mm | 27 ±15 | 24 ±11 |
| Mean stent diameter, mm | 2.9 ±0.47 | 2.8 ±0.41 |
| Maximum pressure, atm | 14 ±3.5 | 14 ±4.1 |
| Treatment of a bifurcation, n (%) | 216 (15%) | 14 (17%) |
| Overlapping stents, n (%) | 329 (22%) | 13 (16%) |
| Lesion complexity | | |
| A | 151 (10%) | 16 (20%) |
| B1 | 491 (33%) | 20 (24%) |
| B2 | 180 (12%) | 9 (11%) |
| C | 654 (44%) | 37 (45%) |

Values are n (%), mean ± standard deviation, or n (%). CABG = coronary artery bypass grafting, DES = drug eluting stent, PCI = percutaneous coronary intervention. *These were lesions in which a staged PCI was planned according to the definition, but which were in the end revascularized by CABG.

**Figure S1. Primary Endpoint Events According to QFR**

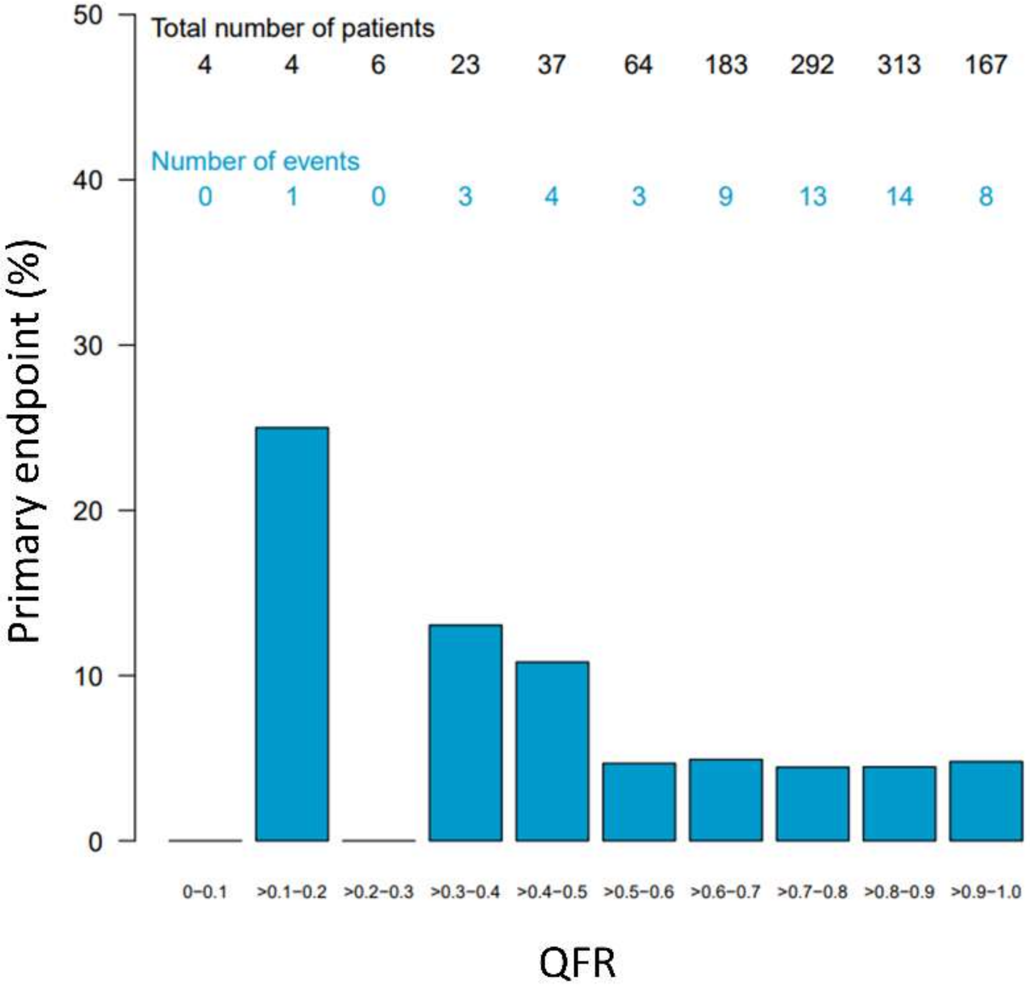


Percentage (%) of primary endpoint events according to Quantitative Flow Ratio (QFR).
